# Reward-Guided Generation Improves the Scientific Utility of Synthetic Biomedical Data

**DOI:** 10.64898/2026.03.11.26348077

**Authors:** Nicholas J. Jackson, Natalia Espinosa-Dice, Chao Yan, Bradley A. Malin

## Abstract

Synthetic data generation is a promising approach for biomedical data sharing and dataset augmentation, yet existing methods lack mechanisms to preserve statistical properties necessary for scientific analysis. To address this, we introduce RLSYN+REG, a reinforcement learning-driven generative model, which encourages that regression models trained on synthetic data reproduce the coefficients and predictions of their real-data counterparts. We evaluate RL-Syn+Reg on MIMIC-III and the American Community Survey (ACS) across regression model reproduction, fidelity to real data, and privacy. Synthetic data from RLSyn+Reg substantially improves upon that of RLSyn, raising correlations between real and synthetic regression coefficients from 0.054 to 0.600 on MIMIC-III and from 0.160 to 0.376 on ACS. Predictive performance also improves, reducing the gap between real-data baselines by 81.4% and 97.6% on MIMIC-III and ACS, respectively. These improvements come with negligible cost to fidelity or privacy and are robust to reductions in training data.

## Introduction

Synthetic data generation has emerged as a practical tool for addressing several needs in biomedical research, including sharing data under privacy constraints and augmenting datasets with limited amounts of records on rare or underrepresented subgroups. The first of these challenges arises due to the limitations in patient-level data sharing imposed by various regulatory frameworks (e.g., the Privacy Rule of the Health Insurance Portability and Accountability Act), which leads to data silos that limit reproducibility and large-scale collaboration^1^. Synthetic data can address this problem by emulating the statistical patterns of real datasets while removing direct connections to real individuals, enabling researchers to share and collaborate more openly, train and validate machine learning models, and perform analyses that would otherwise be infeasible^2^. At the same time, synthetic data offers the ability to augment existing datasets by generating additional samples for underrepresented or rare subgroups^3^. By deliberately oversampling such populations, researchers can, in principle, mitigate the effects of small sample sizes or non-representative datasets that would otherwise render analyses statistically underpowered^4^ or lead to biased machine learning^5^ and regression models^6^. Together, these use cases have driven growing interest in synthetic data as a practical tool for advancing biomedical research^7–9^. Yet the value of synthetic data heavily depends on whether it preserves the specific statistical relationships that researchers care about. A synthetic dataset that distorts associations between clinical variables, for example, by attenuating the relationship between an intervention and mortality, or misrepresenting the prevalence of a disease subgroup, can yield misleading conclusions that undermine the very research it was meant to enable^10^.

Despite these concerns, existing methods for synthetic data generation optimize for general measures of statistical fidelity, but seldom provide researchers with the means to specify or enforce particular desired properties in the synthetic data. Recent work has considered the application of constrained generative models to address this problem. However, such approaches are limited in several ways. First, such constraints are often restricted to simple rules, such as ordinal relationships between clinical measurements (e.g., the feature representing the minimum value of a vital sign must not exceed the median of that vital sign)^11^. Second, other methods require embedding the knowledge of a causal graph in generative models^12^, which may be difficult to satisfy in research datasets where researchers are still trying to establish such relationships.

Reinforcement learning (RL) offers a natural framework for addressing these limitations. Unlike standard generative training objectives, RL reward functions can encode researcher-specified objectives, such as preserving demographic proportions or maintaining associations between clinical variables and outcomes, without requiring those objectives to be differentiable. This means that a single generative architecture can be adapted to a wide range of generation tasks simply by modifying the reward signal, with no changes to the underlying model.

In this study, we extend RLSyn^13^, a RL-based generative model, by introducing a regression-based reward that enforces that regression models trained on synthetic data reproduce the coefficients and predictions of their real-data counterparts. The resulting model, which we refer to as RLSyn+Reg, steers the generator toward synthetic data that preserves the statistical relationships present in the real data. We demonstrate the efficacy of this approach on two regression tasks using tabular biomedical datasets: the development of clinical risk scores and the analysis of socioeconomic patterns across demographic subgroups. We evaluate RLSyn+Reg on a dataset of ICU admissions from the MIMIC-III critical care database^14^ and a dataset of respondents from the American Community Survey (ACS)^15^, a survey capturing demographic and socioeconomic features often linked to clinical data as social determinants of health. For each dataset, we assess the resulting synthetic data across three axes of performance: i) fidelity to the real data distribution, ii) privacy risk to participants, and iii) scientific utility, measured by the degree to which the regression model fit on synthetic data aligns with the real-data regression. Given that synthetic data is most valuable in data-scarce settings, we additionally examine robustness to reductions in training data size.

To the best of our knowledge, this is the first study to demonstrate that targeted RL reward functions can improve the scientific utility of synthetic biomedical data. We show that RLSyn+Reg meaningfully improves coefficient recovery (i.e., the agreement between regression coefficients estimated on synthetic versus real data) and predictive performance with only minor reductions in distributional fidelity and no measurable impact on privacy.

## Methods

### Datasets

We derive our first dataset from MIMIC-III^14^, a publicly available critical care database containing records from Beth Israel Deaconess Medical Center. Our cohort comprises the 27,594 ICU admissions who spent at least 24 hours in the ICU. We include continuous variables that were recorded in at least 20% of all visits, yielding 66 continuous variables. These variables include vital signs (e.g., heart rate and oxygen saturation) and laboratory values (e.g., glucose and white blood cell count). Additionally, we include four binary intervention indicators capturing 1) mechanical ventilation, 2) vasopressor administration, 3) administration of a crystalloid bolus, and 4) the use of non-invasive ventilation. Lastly, we include patient-level features including gender, age, race, insurance status (grouped into private, public, or uninsured), admission type (elective, emergency, or urgent), as well as whether the patient died during their hospital stay. Missing values for continuous variables are imputed using chained multiple imputation^16^.

The second dataset is derived from the American Community Survey from Tennessee in 2018^15^. This data is commonly used in epidemiological studies to incorporate non-clinical factors potentially affecting health outcomes. In this study, we study socioeconomic predictors of public income assistance in 54,452 survey participants. We include age, years of school, marital status, sex, disability status, 1-year mobility (i.e., whether they had moved in the last year), military/veteran status, whether they had ever been institutionalized, the amount of income assistance they receive, whether they have health insurance, their race, and their citizenship status.

Table 1 summarizes the baseline characteristics of both cohorts, stratified by outcome group. The substantial differences between these groups in their demographic and clinical distributions underscore the importance of preserving these associations in synthetic data, as failing to do so would undermine the validity of downstream analyses. Continuous values for both datasets are normalized to have zero-mean and unit standard deviation before generation.

**Table 1.** Baseline characteristics of MIMIC-III and ACS cohorts, stratified by regression outcome (in-hospital mortality for MIMIC-III; receipt of public income assistance for ACS). For descriptive purposes, public income assistance is binarized as any versus none. P-values are from two-sample t-tests for continuous variables and chi-squared tests for categorical variables, indicating differences in demographics between outcome groups.

|  |  | Overall | Group 1 | Group 2 | P-Value |
| --- | --- | --- | --- | --- | --- |
| Panel A: MIMIC-III |  |  |  |  |  |
|  |  | (n = 27,594) | Survived (n = 25,161) | Died (n = 2,433) |  |
| Age, mean (SD) |  | 62.7 (16.7) | 62.2 (16.8) | 68.6 (15.1) | <0.001 |
| Gender | Female | 11,579 (42.0%) | 10,471 (41.6%) | 1,108 (45.5%) | <0.001 |
|  | Male | 16,015 (58.0%) | 14,690 (58.4%) | 1,325 (54.5%) |  |
| Race | Black/African American | 2,147 (7.8%) | 2,010 (8.0%) | 137 (5.6%) | <0.001 |
|  | Other | 5,907 (21.4%) | 5,245 (20.8%) | 662 (27.2%) |  |
|  | White | 19,540 (70.8%) | 17,906 (71.2%) | 1,634 (67.2%) |  |
| Insurance | Private | 9,885 (35.8%) | 9,276 (36.9%) | 609 (25.0%) | <0.001 |
|  | Public | 17,357 (62.9%) | 15,574 (61.9%) | 1,783 (73.3%) |  |
|  | Uninsured | 352 (1.3%) | 311 (1.2%) | 41 (1.7%) |  |
| Admission Type | Elective | 4,925 (17.8%) | 4,828 (19.2%) | 97 (4.0%) | <0.001 |
|  | Emergency | 21,906 (79.4%) | 19,650 (78.1%) | 2,256 (92.7%) |  |
|  | Urgent | 763 (2.8%) | 683 (2.7%) | 80 (3.3%) |  |
| Panel B: ACS |  |  |  |  |  |
|  |  | (n = 54,452) | No Assistance (n = 53,846) | Assistance (n = 606) |  |
| Age, mean (SD) |  | 49.1 (18.7) | 49.2 (18.7) | 46.6 (17.3) | <0.001 |
| Gender | Female | 28,235 (51.9%) | 27,814 (51.7%) | 421 (69.5%) | <0.001 |
|  | Male | 26,217 (48.1%) | 26,032 (48.3%) | 185 (30.5%) |  |
| Race | Asian | 883 (1.6%) | 881 (1.6%) | 2 (0.3%) | <0.001 |
|  | Black/African American | 6,628 (12.2%) | 6,523 (12.1%) | 105 (17.3%) |  |
|  | Other | 663 (1.2%) | 650 (1.2%) | 13 (2.1%) |  |
|  | Two or More | 735 (1.3%) | 721 (1.3%) | 14 (2.3%) |  |
|  | White | 45,543 (83.6%) | 45,071 (83.7%) | 472 (77.9%) |  |

### Generative Model

We build on the RLSYN framework^13^, which reformulates the training of a Generative Adversarial Network (GAN) as an RL problem. In this framework, a generator acts as a policy network that learns to produce realistic synthetic data, while a discriminator serves as a critic, providing a reward signal to guide generation. The generator is a multi-layer perceptron that maps a random noise vector to a synthetic row via separate output heads for continuous, binary, and categorical features. Continuous features, such as heart rate, age, and oxygen saturation, are modeled as Normal distributions with learned means and variances. Similarly, binary features, such as vasopressor administration and ventilator usage, are modeled as Bernoulli distributions with learned success probabilities and categorical features, such as race and insurance status, are modeled as categorical distributions with learned class probabilities. Rather than producing deterministic outputs, the generator samples from these distributions at each iteration, enabling the model to track and adjust the probability it assigns to each generated sample. The discriminator is a multi-layer perceptron that receives a tabular row and outputs a scalar realism score relative to the training data..

### Training Procedure

The generator and discriminator are trained in an alternating fashion using Proximal Policy Optimization (PPO)^17^. At each iteration, the generator produces a batch of synthetic rows, after which the discriminator scores each row, and the generator is then updated to produce rows that received higher scores. The generator is updated using the standard PPO clipped surrogate objective and the discriminator is updated via the standard discriminator loss for a GAN (i.e., it is trained using binary cross-entropy to distinguish real from synthetic data). The discriminator loss also incorporates an R1 gradient penalty, which encourages the discriminator to learn a smooth decision boundary between real and synthetic data, providing a more stable reward signal for generator updates. The weighting coefficient for the R1 penalty is selected via hyperparameter search.

Unlike a standard GAN, where the generator must backpropagate through the discriminator, PPO treats the discriminator score as a black-box reward signal, decoupling the generator and discriminator. The efficiency of this approach was found to outperform diffusion- and GAN-based tabular data generation^13^. More centrally to this work, this decoupling enables the use of flexible reward functions, without any changes to the generator architecture.

### Regression-Based Reward

We extend RLSYN with a regression-based auxiliary reward, yielding a model we refer to as RLSYN+REG, which penalizes synthetic rows whose outcome distribution is inconsistent with the regression structure observed in real data. Specifically, we fit a regression model *f* on the real training data prior to training and use the learned coefficients to define a fixed scoring function for each generated row *x*.

The architecture of RLSYN provides direct access to the generator’s conditional distribution over the outcome (*q*(*x*) ≜ *p*(*y* = 1 | *x*)) prior to sampling. That is, for binary outcomes, the activation of the output head before the Bernoulli draw, and for continuous outcomes, the conditional mean of the Gaussian policy head before sampling. This allows us to directly penalize the generator’s conditional outcome probability *q*(*x*) against the regression model’s predicted probability *f* (*x*). For both continuous and binary outcomes, the per-sample penalty takes the form:

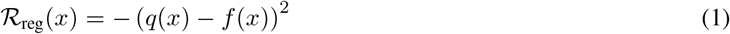

For example, if the generative model believes that a generated record *x* has a mortality probability of 40% (*q*(*x*) = 0.4), but the regression model predicts a mortality probability of 70% (*f* (*x*) = 0.7), then the generator will receive the reward− (0.4 −0.7)^2^. This reward encourages the generator to push *q*(*x*) closer to the fixed value *f* (*x*). This penalty is introduced at iteration *t*′ and increased linearly from 0 to its full weight *λ*. Both *λ* and *t*′ are hyperparameters selected via Optuna. The total per-sample reward (for a sample *x*) at iteration *t* is then:

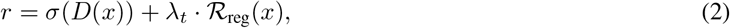

where *σ*(*D*(*x*)) is the realism score of the discriminator *D* on the synthetic data *x*. This reward replaces the discriminator-only reward in the base RLSyn training procedure and is used to compute the generator loss via PPO.

### Evaluation

We evaluate synthetic data across three performance axes: 1) downstream utility, 2) distributional fidelity, and 3) participant privacy. Our primary analysis compares data generated by the default RLSyn to that generated by RLSyn+Reg. Since the settings where synthetic data are most valuable (e.g., research on rare diseases or health disparities) are often characterized by limited training data, we additionally perform a sensitivity analysis in which we vary the size of the training dataset to assess whether the benefits of RLSyn+Reg are robust in low-data settings. All experiments are repeated across three random seeds and results are averaged across runs.

Hyperparameters are selected via Bayesian search^18^ over 10,000 training steps, with the final models trained for 50,000 and 25,000 steps on MIMIC and ACS, respectively. For RLSyn, we tune the number of PPO updates, the ratio of discriminator to generator updates, the R1 penalty weight, and the learning rates, depth, and width of both networks. For the regression-based reward, a secondary search tunes the regression penalty weight *λ* and the start iteration for the regression penalty *t*′, selecting the best tradeoff between regression performance and overall data quality.

### Utility

We assess utility by measuring the preservation of scientifically meaningful relationships in the synthetic data. Specifically, we fit regression models on real and synthetic datasets and compare their coefficients and held-out predictive performance. For the MIMIC dataset, we fit a logistic regression model to predict mortality with features spanning: 1) respiratory status (oxygen saturation, mechanical ventilation), 2) cardiovascular function (mean arterial pressure), 3) vasopressor use (lactate), 4) renal function (creatinine), 5) coagulation (platelets), 6) hepatic function (bilirubin), 7) neurological status (Glasgow Coma Scale), and 8) demographics (age, sex, race). For the ACS dataset, we fit an ordinary least-squares (OLS) regression model to predict public income assistance as a function of age, years of education, sex, and race. These analyses represent clinically and socially meaningful applications of synthetic data: preserving the relationships between physiological deterioration and mortality in critical care, and the socioeconomic patterns that underpin health disparities research. We also measure coefficient correlation — correlation between real and synthetic model coefficients. Performance of the regression models on MIMIC and ACS are assessed via area under the receiver operating characteristic curve (AUC) and root-mean squared error (RMSE), respectively.

### Fidelity

We measure distributional fidelity according to two criteria. The first is Dimension-Wise Difference (DWD)^19^, which captures univariate similarity: absolute prevalence difference (APD) for binary and categorical variables, and Wasserstein distance for continuous variables (normalized to [0, 1]), averaged across all features. The second is Column-wise Correlation (CWC) difference^20^, which captures multivariate structure as the mean absolute difference between the real and synthetic feature correlation matrices (normalized by number of correlations). Lower values indicate higher fidelity for both metrics.

### Privacy

We assess privacy via membership inference risk, a widely used measure of synthetic data privacy^21,22^. While other privacy measures exist^20,23^, membership inference is a necessary prerequisite: if an adversary cannot identify whether an individual was in the training data, no further claims about the privacy of that individual can be made. Specifically, we simulate a scenario in which an adversary has access to 5,000 real data records, 2,500 that were used to train a generator and 2,500 that were not. The adversary does not know which records were used in training and attempts to infer training membership based on each record’s nearest-neighbor distance to the synthetic data. AUC of the attack is computed by aggregating over classification thresholds, with values near 0.5 indicating that the attack success is near random-chance and the synthetic data do not meaningfully leak information about training records.

Code to reproduce all analyses is available at https://github.com/nicholas-j-jackson/RL-Guided-Generation.

## Results

We organize our results in two parts. We begin with our experimental results, demonstrating that RLSyn+Reg improves regression coefficient recovery and predictive performance on MIMIC-III and ACS, and that these improvements are robust to reductions in training data size. We then provide a brief theoretical analysis establishing that these improvements are not an artifact of the specific datasets or outcomes studied here, but follow from general properties of the regression-based reward, supporting the generalizability of the approach.

### Experimental Results

Table 2 reports fidelity, utility, and privacy metrics for RLSyn and RLSyn+Reg across both datasets. The fidelity of RLSyn+Reg is high, as evidenced by close alignment between real and synthetic feature prevalences and pairwise correlations across both datasets (Figure 1). Privacy is similarly well-preserved, with MIA AUC near 0.5 across both models and datasets, indicating that the synthetic data do not meaningfully leak information about individual training records.

**Table 2.**
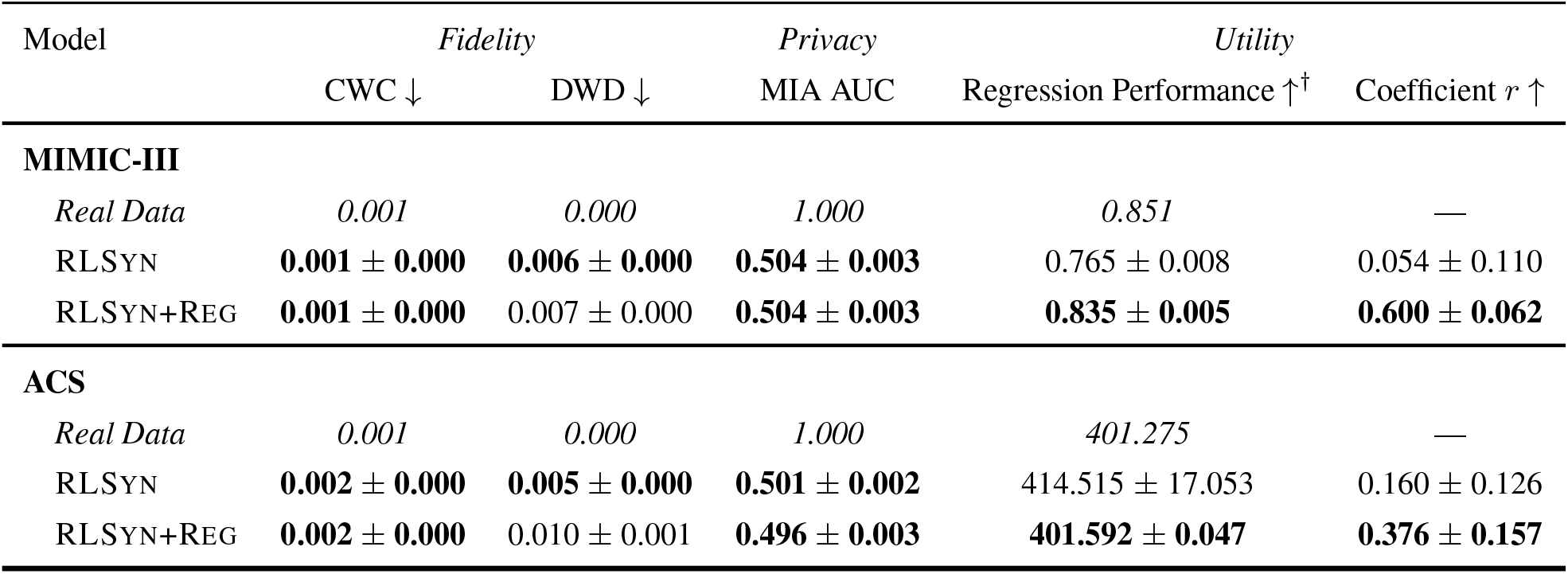
Effect of Regression Reward. ^†^Regression performance is measured via AUC (↑) for MIMIC-III and RMSE (↓) for ACS. Means ± standard deviations are computed across three training runs. *Real data* results for CWC and DWD were computed between the training data and test data. The best results for each column are bolded.

| Model | Fidelity |  | Privacy | Utility |  |
| --- | --- | --- | --- | --- | --- |
| | CWC ↓ | DWD ↓ | MIA AUC | Regression Performance $\uparrow^{\dagger}$ | Coefficient $r$ ↑ |
| **MIMIC-III** |  |  |  |  |  |
| *Real Data* | 0.001 | 0.000 | 1.000 | 0.851 | — |
| RLSYN | **0.001 ± 0.000** | **0.006 ± 0.000** | **0.504 ± 0.003** | 0.765 ± 0.008 | 0.054 ± 0.110 |
| RLSYN+REG | **0.001 ± 0.000** | 0.007 ± 0.000 | **0.504 ± 0.003** | **0.835 ± 0.005** | **0.600 ± 0.062** |
| **ACS** |  |  |  |  |  |
| *Real Data* | 0.001 | 0.000 | 1.000 | 401.275 | — |
| RLSYN | **0.002 ± 0.000** | **0.005 ± 0.000** | **0.501 ± 0.002** | 414.515 ± 17.053 | 0.160 ± 0.126 |
| RLSYN+REG | **0.002 ± 0.000** | 0.010 ± 0.001 | **0.496 ± 0.003** | **401.592 ± 0.047** | **0.376 ± 0.157** |

**Figure 1:**
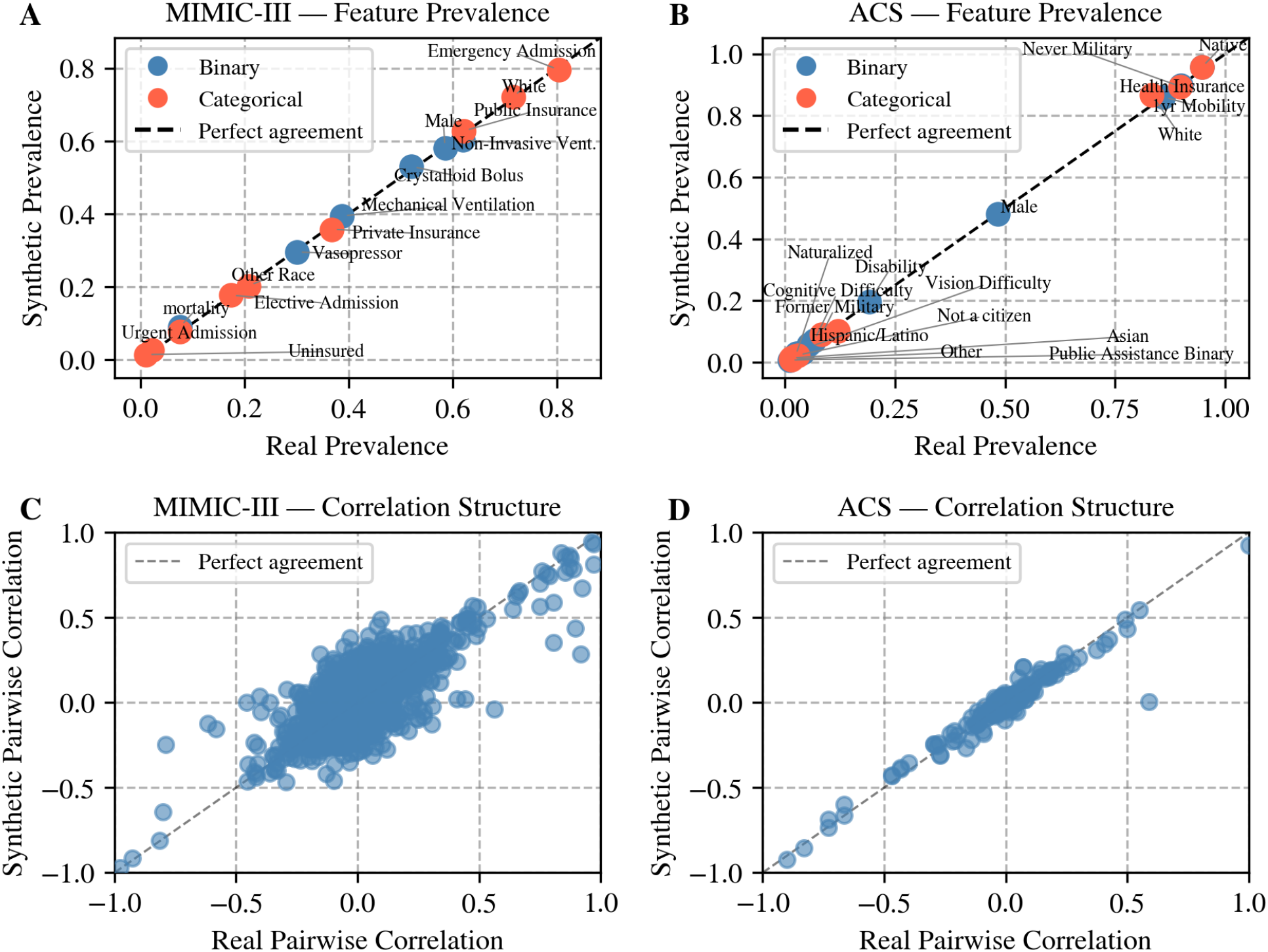
Fidelity of synthetic data generated by RLSyn+Reg across MIMIC-III and ACS. Real (**A**) vs. synthetic feature prevalences for binary (blue) and categorical (orange) features. Points along the dashed diagonal indicate perfect agreement. Pairwise correlations between continuous features in real (**C**) vs. synthetic (**D**) data. Each point represents one feature pair; points along the diagonal indicate perfect preservation of correlation structure.

Despite strong fidelity, base RLSyn preserves key scientific relationships poorly. Notably, regression model performance is low relative to the real data baseline (Table 2: AUC 0.765 vs. 0.851 for MIMIC-III; RMSE 414.515 vs. 401.592 for ACS). Additionally, coefficient *r* is near zero for both datasets (0.054 and 0.160 for MIMIC and ACS, respectively), indicating that base RLSyn does not reliably reproduce the clinically and socioeconomically meaningful associations present in the real data.

Our results further indicate that introducing the regression penalty substantially improves scientific utility, as assessed by regression model performance (Table 2). On MIMIC-III, AUC increases from 0.765 to 0.835, recovering nearly the full gap between RLSyn and the real data baseline. Additionally, coefficient correlation improves from 0.054 to 0.600. Similar results are observed in the ACS dataset, where RMSE improves from 414.515 to 401.592, matching the real data baseline of 401.275 almost exactly, and coefficient correlation increases from 0.160 to 0.376. However, these gains come at a modest cost to fidelity: CWC increases by roughly 7% on MIMIC-III and 24% on ACS, and while DWD scores nearly double, values remain small in absolute terms. Critically, the regression penalty has no measurable impact on privacy risk; MIA AUC remains near 0.5 for RLSyn+Reg across both datasets, indicating that the improved scientific utility is not achieved at the expense of participant privacy.

Figure 2 provides a sensitivity analysis of the regression performance as a function of the training data size. On MIMIC-III, both models degrade as the amount of training data decreases, as expected. However, RLSyn+Reg maintains consistently higher AUC across all fractions, suggesting that the regression penalty provides a stable benefit regardless of sample size. On ACS, regression RMSE is largely insensitive to training data fraction for both models, with RLSyn+Reg matching the real data baseline across all conditions. Together, these results suggest that the benefit of the regression penalty is robust to data scarcity, and does not depend on large sample sizes to take effect.

**Figure 2:**
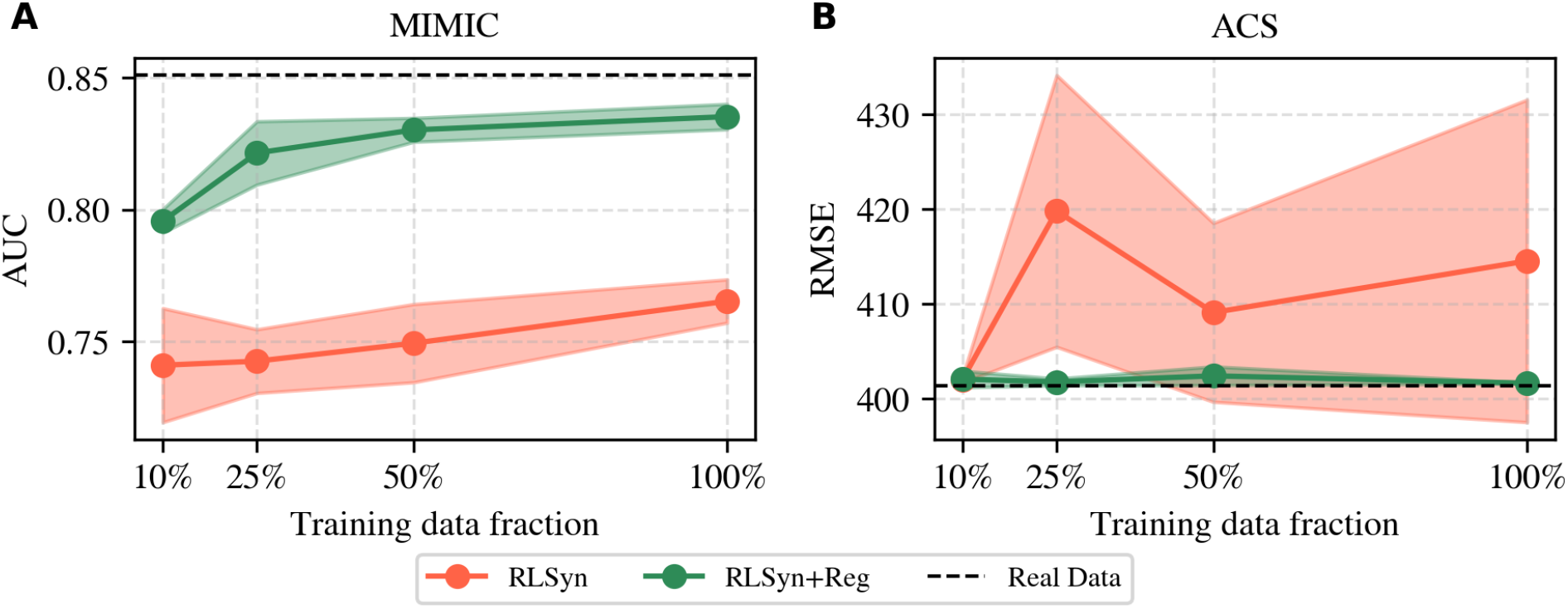
Effect of training data size on regression performance for RLSyn and RLSyn+Reg across MIMIC-III and ACS. **(A)** Regression model AUC on MIMIC-III and **(B)** Regression model RMSE on ACS, both as a function of training data fraction. The dashed horizontal line indicates the real data baseline. Shaded regions denote ± 1 standard deviation across random seeds.

### Theoretical Results

In the proof below, we show that two conditions that are enforced during training (non-degeneracy and conditional probability matching) are sufficient to guarantee that fitting a regression model on the synthetic data recovers the same coefficients as a model fit on the real data. This result holds for both logistic and linear regression, covering the clinical risk score and health disparities settings that we evaluate in the previous section.

Let **x** ∈ ℝ^*d*^ denote a feature vector, *y* ∈ {0,1} a binary outcome, and 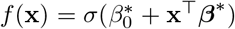 the regression model’s predicted probability, where *σ*(*t*) = (1 + *e*^−*t*^)^−1^ and 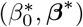 is the maximum likelihood estimate on real data, which uniquely minimizes the strictly convex negative log-likelihood^24^ and satisfies:

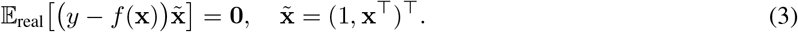

The proof relies on two conditions. First, the synthetic data has sufficient coverage of the feature space. That is, no feature is constant or an exact linear combination of the others (Condition 1: Non-degeneracy). This condition is encouraged by the discriminator, which penalizes synthetic data that does not reflect the variation present in the real data. Second, the generator’s predicted outcome probability *q*(**x**) matches the regression model’s predicted probability *f* (**x**) (Condition 2: Probability Matching), a condition enforced in expectation by minimizing Equation 1.

#### Theorem 1

(Coefficient Recovery). *Under Conditions 1 and 2, fitting logistic regression on synthetic data recovers coefficients* 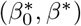*as the maximum likelihood estimate*.

*Proof*. We show that the optimality condition holds on the synthetic distribution:

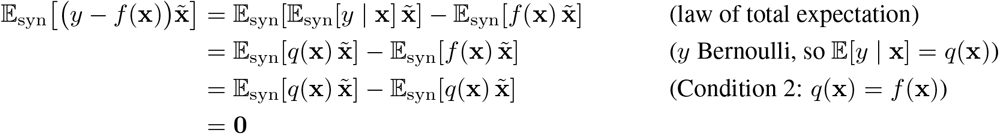

Hence 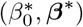 is a stationary point of the synthetic negative log-likelihood. Since the negative log-likelihood is strictly convex under full-rank features (Condition 1), any stationary point is the unique global minimum and therefore the unique maximum likelihood estimate^24^.

The result extends naturally to the continuous outcome setting. For OLS regression, the first-order condition is 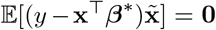. An identical argument shows that aligning the generator’s conditional mean *µ*_*θ*_(**x**) — the mean of the Gaussian policy head — with the OLS prediction *f* (**x**) = **x**^T^***β***^*^ recovers ***β***^*^ on the synthetic data.

In practice, Conditions 1 and 2 likely only hold approximately, and the degree to which coefficients are recovered depends on the quality of the generator’s distributional fit and the strength of the regression penalty. The empirical results above suggest that, despite these approximations, the regression penalty provides meaningful improvements.

## Discussion

Our results demonstrate that RLSyn+Reg, which introduces a regression-based auxiliary reward optimized via reinforcement learning, can guide a generative model to preserve specific statistical relationships present in real data without requiring task-specific architectural changes. While other work has investigated RL to make synthetic data generation more efficient^13^ or performant^25^, the results presented here suggest that reward-guided generation is a promising and underexplored direction for improving the scientific utility of synthetic biomedical data. More specifically, using RLSyn+Reg, a researcher could share a synthetic dataset that maintains the regression relationships observed in a published study on clinical risk scores, allowing others to verify findings or apply the same analytical pipeline without ever accessing the original patient-level data. Similarly, RLSyn+Reg could support a health disparities analysis in a population where direct data access is restricted by regulatory constraints. The sensitivity analysis further demonstrates that these benefits extend to small training datasets, where augmenting datasets with synthetic data is most promising^3,4^.

Importantly, while these results are achieved with no change in privacy risk, we observe small decreases in statistical fidelity. Specifically, the distributions of individual features and the correlations between feature pairs (as measured by DWD and CWC, respectively) are slightly less well-preserved in RLSyn+Reg compared to base RLSyn. This is an expected consequence of steering the generator toward a specific statistical target, as this orients the model’s learning capacity toward coefficient recovery rather than the full data distribution. While this tension is likely unavoidable, a key contribution of our approach is that it is controllable: the penalty weight *λ* and anneal start *t*′ together allow researchers to select the appropriate tradeoff for their use case. For example, a researcher sharing a synthetic ICU dataset to support replication of a published mortality prediction model may accept a larger decrease in fidelity, since the primary goal is preserving the regression relationships rather than the full data distribution. By contrast, a researcher generating synthetic data to augment the training of a machine learning model, where exposure to diverse and realistic feature combinations is essential, may prefer to minimize fidelity costs even at some cost to coefficient recovery.

In addition to the regression setting evaluated in this study, the flexibility of RLSyn+Reg allows it to extend to many other applications. Specifically, because PPO treats the reward as a black-box signal, any computable objective can be introduced as an auxiliary reward without modifying the generator. For researchers, this means that objectives such as preserving the odds ratio for a known risk factor, maintaining the calibration of a clinical prediction model, or reproducing the demographic composition of a patient subgroup can be encoded directly into the generative process without requiring expertise in deep learning or generative modeling. This modularity suggests a broader paradigm of objective-driven synthetic data generation, in which the scientific goals of the end user are encoded directly into the training signal. The results presented here serve as a proof-of-concept of this paradigm in the context of coefficient recovery, and we discuss several promising extensions below.

### Limitations and Future Work

There are several main limitations of this investigation that offer natural extensions into future research. First, our evaluation was limited to tabular data. Tabular data affords clean separation of features into distinct types, and each feature has a direct and measurable impact on downstream statistical analyses, making it a natural starting point for reward-guided generation. However, the generalizability of this approach to other biomedical data modalities (e.g., longitudinal medical records and medical imaging) remains an open question, as the relationship between generated data and downstream statistical properties is less direct in these settings. Future work should explore how regression-based and other targeted rewards can be adapted to these more complex modalities.

Second, our approach applies only to regression models at the population level. However, a key challenge in synthetic data generation is preserving relationships within clinically- or socially-meaningful subgroups. Subgroup-specific reward functions offer an opportunity to improve synthetic data utility for underrepresented groups. For example, by penalizing regression patterns within minority subgroups more heavily than in the overall population, researchers could generate synthetic data that more faithfully reproduces health disparities or differential treatment effects that would otherwise be obscured by population-level optimization. Future work should investigate the use of subgroup-targeted reward functions for fairness-aware synthetic data generation.

Lastly, the rewards we develop aim to replicate relationships that are already present in the real data. A natural extension would be to introduce rewards that either remove undesirable relationships present in the real data, such as biases, or introduce desirable patterns that are absent from it. This would be more challenging than the present work because the discriminator is trained to make the synthetic data resemble the real data as closely as possible, which means that it would actively work against a reward that pushes the generator away from the real data distribution. Balancing these competing signals would require careful reward design and weighting. However, if successful, such a method could be used to penalize associations between protected attributes and biased measures of clinical outcomes^26^, or to incorporate clinical knowledge about the underlying causal structure of highly-confounded observational datasets^12^.

## Conclusion

Existing approaches to synthetic data generation optimize for general distributional fidelity without providing researchers any means to enforce the specific statistical properties their analyses depend on. We introduce RLSyn+Reg, which addresses this gap by augmenting a reinforcement learning-based generative model with a regression-based auxiliary reward. Across two tabular datasets spanning clinical and social scientific applications, RLSyn+Reg substantially improves coefficient recovery and predictive performance of regression models fit on synthetic data, with only minor reductions in fidelity and no measurable impact on privacy. These benefits are robust to reductions in training data size, suggesting that RLSyn+Reg remains effective in the data-scarce settings where synthetic data is most needed. As synthetic data becomes increasingly central to reproducible and privacy-preserving biomedical research, methods that allow researchers to specify and enforce meaningful statistical properties will be critical to its adoption in practice. The modularity of the reward-guided framework, in which any computable objective can be encoded as an auxiliary reward without architectural changes, positions it as a promising foundation for future work on objective-driven synthetic data generation.

## Data Availability

All data used in the present study are available online. Complete instructions are available on github: https://github.com/nicholas-j-jackson/RL-Guided-Generation

## Acknowledgments

This work was supported by the National Library of Medicine (T15LM007450, K99LM014428) and the National Human Genome Research Institute (U54HG012510).

## References

1. Schreiber R, Koppel R, Kaplan B. What Do We Mean by Sharing of Patient Data? DaSH: A Data Sharing Hierarchy of Privacy and Ethical Challenges. Applied Clinical Informatics. 2024 Oct;15(05):833–41. Available from: http://www.thieme-connect.de/DOI/DOI?10.1055/a-2373-3291.

2. Giuffrè M, Shung DL. Harnessing the power of synthetic data in healthcare: innovation, application, and privacy. npj Digital Medicine. 2023 Oct;6(1):186. Available from: https://www.nature.com/articles/s41746-023-00927-3.

3. Sagers LW, Diao JA, Melas-Kyriazi L, Groh M, Rajpurkar P, Adamson AS, et al. Augmenting medical image classifiers with synthetic data from latent diffusion models. arXiv; 2023. ArXiv:2308.12453 [cs]. Available from: http://arxiv.org/abs/2308.12453.

4. Wang D, Florian H, Lynch S, Robieson W, Zhuang R, Kusiak C, et al. Using AI-generated digital twins to boost clinical trial efficiency in Alzheimer’s disease. Alzheimer’s & Dementia: Translational Research & Clinical Interventions. 2025 Oct;11(4):e70181. Available from: https://alz-journals.onlinelibrary.wiley.com/doi/10.1002/trc2.70181.

5. Jackson NJ, Yan C, Malin BA. Enhancement of Fairness in AI for Chest X-ray Classification. AMIA Annual Symposium proceedings AMIA Symposium. 2024;2024:551–60. Available from: https://pmc.ncbi.nlm.nih.gov/articles/PMC12099404/.

6. Juwara L, El-Hussuna A, El Emam K. An evaluation of synthetic data augmentation for mitigating covariate bias in health data. Patterns. 2024 Apr;5(4):100946. Available from: https://linkinghub.elsevier.com/retrieve/pii/S266638992400045X.

7. Abul-Husn NS, Kenny EE. Personalized Medicine and the Power of Electronic Health Records. Cell. 2019 Mar;177(1):58–69. Available from: https://linkinghub.elsevier.com/retrieve/pii/S0092867419302223.

8. Perlman SE, McVeigh KH, Thorpe LE, Jacobson L, Greene CM, Gwynn RC. Innovations in Population Health Surveillance: Using Electronic Health Records for Chronic Disease Surveillance. American Journal of Public Health. 2017 Jun;107(6):853–7. Available from: https://ajph.aphapublications.org/doi/full/10.2105/AJPH.2017. 303813.

9. Sadler MC, Apostolov A, Cevallos C, Auwerx C, Ribeiro DM, Altman RB, et al. Leveraging large-scale biobank EHRs to enhance pharmacogenetics of cardiometabolic disease medications. Nature Communications. 2025 Mar;16(1):2913. Available from: https://www.nature.com/articles/s41467-025-58152-3.

10. Koul A, Duran D, Hernandez-Boussard T. Synthetic data, synthetic trust: navigating data challenges in the digital revolution. The Lancet Digital Health. 2025 Dec:100924. Available from: https://linkinghub.elsevier.com/retrieve/pii/S2589750025001062.

11. Yan C, Zhang Z, Nyemba S, Malin BA. Generating Electronic Health Records with Multiple Data Types and Constraints. AMIA Annual Symposium proceedings AMIA Symposium. 2020;2020:1335–44.

12. Asiaee A, Liang ZJ, Yan C. CausalWrap: Model-Agnostic Causal Constraint Wrappers for Tabular Synthetic Data. arXiv; 2026. ArXiv:2603.02015 [cs]. Available from: http://arxiv.org/abs/2603.02015.

13. Espinosa-Dice N, Jackson NJ, Yan C, Lee A, Malin BA. A Reinforcement Learning Approach to Synthetic Data Generation. arXiv; 2025. ArXiv:2512.21395 [cs]. Available from: http://arxiv.org/abs/2512.21395.

14. Johnson AEW, Pollard TJ, Shen L, Lehman LwH, Feng M, Ghassemi M, et al. MIMIC-III, a freely accessible critical care database. Scientific Data. 2016 May;3(1):160035. Available from: https://www.nature.com/articles/sdata201635.

15. Ding F, Hardt M, Miller J, Schmidt L. Retiring adult: new datasets for fair machine learning. arXiv; 2022. ArXiv:2108.04884 [cs, stat]. Available from: http://arxiv.org/abs/2108.04884.

16. Buuren SV, Groothuis-Oudshoorn K. mice: Multivariate Imputation by Chained Equations in R. Journal of Statistical Software. 2011;45(3). Available from: http://www.jstatsoft.org/v45/i03/.

17. Schulman J, Wolski F, Dhariwal P, Radford A, Klimov O. Proximal Policy Optimization Algorithms. arXiv; 2017. ArXiv:1707.06347 [cs]. Available from: http://arxiv.org/abs/1707.06347.

18. Akiba T, Sano S, Yanase T, Ohta T, Koyama M. Optuna: A Next-generation Hyperparameter Optimization Framework. arXiv; 2019. ArXiv:1907.10902 [cs]. Available from: http://arxiv.org/abs/1907.10902.

19. Yan C, Yan Y, Wan Z, Zhang Z, Omberg L, Guinney J, et al. A Multifaceted benchmarking of synthetic electronic health record generation models. Nature Communications. 2022 Dec;13(1):7609. Available from: https://www.nature.com/articles/s41467-022-35295-1.

20. Yan C, Zhang Z, Nyemba S, Li Z. Generating Synthetic Electronic Health Record Data Using Generative Adversarial Networks: Tutorial. JMIR AI. 2024 Apr;3:e52615. Available from: https://ai.jmir.org/2024/1/e52615.

21. Zhang Z, Yan C, Malin BA. Membership inference attacks against synthetic health data. Journal of Biomedical Informatics. 2022 Jan;125:103977. Available from: https://linkinghub.elsevier.com/retrieve/pii/S1532046421003063.

22. Breugel Bv, Sun H, Qian Z, Schaar Mvd. Membership Inference Attacks against Synthetic Data through Overfitting Detection. arXiv; 2023. ArXiv:2302.12580 [cs]. Available from: http://arxiv.org/abs/2302.12580.

23. El Emam K, Mosquera L, Bass J. Evaluating Identity Disclosure Risk in Fully Synthetic Health Data: Model Development and Validation. Journal of Medical Internet Research. 2020 Nov;22(11):e23139. Available from: http://www.jmir.org/2020/11/e23139/.

24. Albert A, Anderson JA. On the existence of maximum likelihood estimates in logistic regression models. Biometrika. 1984;71(1):1–10. Available from: https://academic.oup.com/biomet/article-lookup/doi/10.1093/biomet/71.1.1.

25. Theodorou B, Xiao C, Glass L, Sun J. MediSim: Multi-granular simulation for enriching longitudinal, multi-modal electronic health records. Patterns. 2025 Jun;6(6):101261. Available from: https://linkinghub.elsevier.com/retrieve/pii/S2666389925001096.

26. Ganin Y, Ustinova E, Ajakan H, Germain P, Larochelle H, Laviolette F, et al. Domain-Adversarial Training of Neural Networks. Journal of Machine Learning Research. 2016;17(59):1–35. Available from: http://jmlr.org/papers/v17/15-239.html.

